# LITERATUR REVIEW : DEVELOPMENT OF DISASTER EDUCATION IN MULTI AGENT BASED EARLY EMERGENCY HANDLING IN THE PRE-HOSPITALS AREA

**DOI:** 10.1101/2022.01.23.22269252

**Authors:** Priyo Mukti Pribadi Winoto, Lono Wijayanti, Moses Glorino Rumambo Pandin

**Author notes:** Corresponding author: Priyo Mukti Pribadi Winoto, Campus C Mulyorejo Surabaya 60115, Universitas Airlangga.

## Abstract

**Background:** Disaster risk reduction involves several sectors, legal, social, structural, economic, technological, educational, environmental health, with improved preparedness will be able to reduce the impact of disasters. Capacity that continues to be added with knowledge about rapid disaster response to disasters can make resilience dealing with disasters and preventing the emergence of disaster risks.

**Purpose:** Conduct a literature review on articles that examine the importance of disaster education in the initial handling of emergencies in the hospital area

**Design:** Literature review

**Method:** Using databases with electronic search on ProQuest, SAGE, and Science Direct published in 2017-2021

**Results:** 100 articles were used in the review. These articles discuss the importance of disaster education in the initial handling of emergencies in the hospital area. The 15 articles reviewed are original research.

**Conclusion:** Local communities are well placed to play a central role in hazard identification, development of preparedness plans, detection and response to emergencies, and implementation of recovery efforts. Community leaders and local health workers (e.g. family doctors, nurses, midwives, pharmacists, community health workers) can build public trust, disseminate information, and identify people at risk

## INTRODUCTION

Preventing and reducing the number of fatalities from the exposed population is crucial in the context of disaster risk management(Nugrahandika & Putri, 2021). In dealing with disasters, it involves many lines with a combination of lines of Health, economy, technology and institutions that are continuously intervened and treated in disaster preparedness events that trigger community resilience to personal with the end goal of increasing expertise in responding to conditions that occur as a result of disasters (Sendai, 2021).

Improvements in disaster risk management together with improved living standards have significantly reduced mortality rates from natural hazards (UNDRR, 2021). Preparation is the first step to an all-hazards approach to disaster. Mass casualty incident planning includes an integrated disaster plan, a clear chain of command, hazard vulnerability assessment, surge capacity plan, and triage strategy. The hospital will need an emergency management committee or department whose function is to facilitate disaster training and annual disaster drills. Disaster drills that can help prepare for mass critical care (Sendai, 2019). The bottom-up approach includes local coping mechanisms, recognizing them and strengthening community capacities are important in the disaster risk reduction process (Pandey C, 2019)

These types of extreme events result in great morbidity and mortality, especially in low-income countries, as these countries do not have appropriate disaster preparedness and management plans in place. In Nepal, between 1971 and 2016, more than 26,000 natural disasters were reported, which claimed the lives of more than 43,000 Nepalese and left more than 83,000 injured. The trend of catastrophic events has increased drastically since 2000, perhaps as a consequence of the increasing hazards of climate change. On September 11, 2021, there were 1,873 disasters recorded. The dominant natural disasters are floods, followed by extreme weather and landslides. Natural disasters affected and displaced 5,952,018 people, while as many as 511 people died and 70 were missing and 12,892 people were injured. In addition to natural disasters on April 13, 2020, the government declared the spread of COVID-19 as a non-natural national disaster (BNPB, 2021)

Emergency and disaster risk management is everyone’s business (WHO, 2021). Disaster response, as one of the phases of disaster management, is very important to reduce the impact of disasters and increase the resilience of the public sector and society after a disaster occurs (Rahmayanti, 2021). Disaster risks faced by residents in disaster-prone areas need to be reduced by building good ones. Disaster management. Disaster education based on local wisdom is very important to support formal education. Education in schools or in various disaster counseling activities carried out by the government (Septiana, 2019). Local communities are well placed to play a central role in hazard identification, development of preparedness plans, detection and response to emergencies, and implementation of recovery efforts. Community leaders and local health workers (e.g. family doctors, nurses, midwives, pharmacists, community health workers) can build public trust, disseminate information, and identify people at risk (WHO, 2021)

## METHOD

### Search Strategy

The preparation of this literature review uses various databases by conducting electronic searches on ProQuest, SAGE and Science Direct. The search is limited to articles published in the last five years from 2019 to 2021 which are available in English. Several terms or keywords are combined to get the right article as a search strategy such as using the term “disaster education in early emergency management in the hospital area”

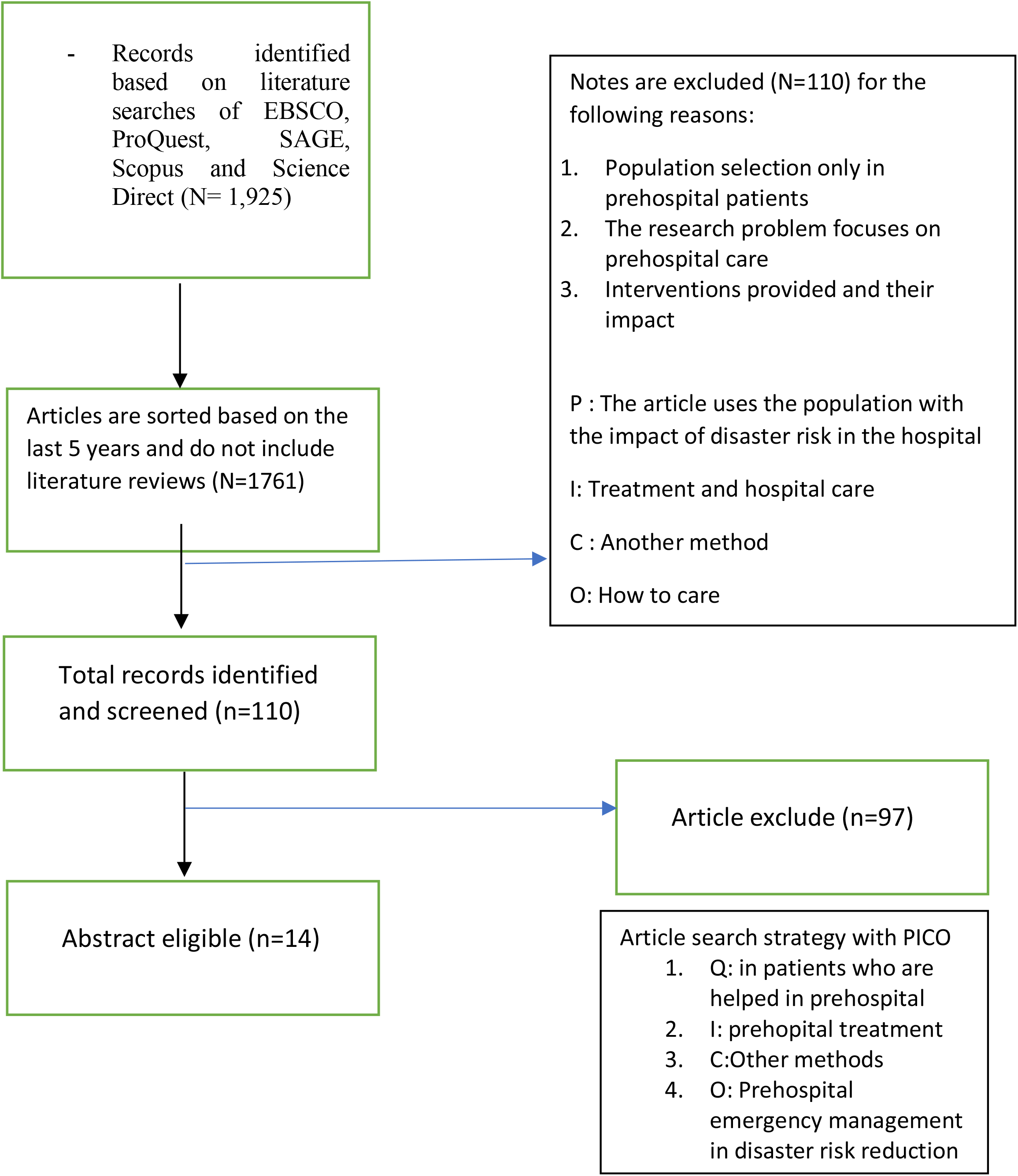

**Table 1.** Results.

| No | Title and Author | Method (Design, Variable, Sample, Instrument) | Results |
| --- | --- | --- | --- |
| 1 | Cpr, P., & Ecc, D. (2020). The main focus of the 2020 American Heart Association Guidelines for CPR and ECC. | **D:** prospective cohort study<br>**S:** 350,000 patients<br>**V:** cpr and ecc Pedoman guidelines<br>**I:** ilcor 2020 | Basic life support can be taught to the community to cope with risks |
| 2 | BNPB. (2020). Guidelines for the Quick Handling of Covid-19 Medical and Public Health in Indonesia. March 23. | **D:** descriptive analytic observational with cross sectional approach.<br>**S:** 14,705 patients<br>**V:** medical prompt treatment guidelines<br>**I:** module | Cardiopulmonary resuscitation targeting lay people |
| 3 | BNPB. (2021). Bencana Alam. | **D:** a cohort study<br>**S:**<br>**V:** bencana Alam<br>**I:** observation checklist | Types of natural disasters |
| 4 | Duong, H. v., Herrera, LN, Moore, JX, Donnelly, J., Jacobson, KE, Carlson, JN, Mann, NC, & Wang, HE (2018). | **D:** compare<br>**S:** 16,116,219 patients<br>**V:** prehospital emergency care.<br>**I:** observation sheet | EMS personnel must be ready to treat patients |
| 5 | Khumairoh, Z., Widana, IDKK, & Sumantri, SH (2021). | **D:** **qualitative description**<br>**S:**<br>**V:** disaster risk reduction in Indonesia<br>**I:** FGD | The results of this study indicate that the role of disaster communication can help the government provide and disseminate information to the public in order to support the success of disaster risk reduction. |
| 6 | Maharani, D., Afifuddin, M., Putri, DA, Setiawan, K., & Ansari, SA (2019). Braille Monopoly Game as a Tool of Disaster Mitigation | **D:** pretest and posttest<br>**S:** **11 patients** | There is an increase in knowledge and awareness of the |
|  | Education for Visual Disabilities. IOP Conference Series. Earth and Environmental Science, 273(1). https://doi.org/http://dx.doi.org/10.1088/1755-1315/273/1/012042 | **V:** disaster mitigation education<br>**I:**monca (disaster monopoly) | importance of disaster mitigation. |
| 7 | Merchant, RM, Topjian, AA, Panchal, AR, Cheng, A., Aziz, K., Berg, KM, Lavonas, EJ, & Magid, DJ (2020). | **D:** observation<br>**S:** 12 500patient<br>**V:** 2020 AHA guidelines<br>**I:**training | Most of the variation in survival rates is thought to be due to the activation of the Chain of Survival, critical actions that must occur sequentially to maximize the chances of survival. |
| 8 | Morrow, KE (2020). Development disasters: accountability and remedy in international development projects. American University Law Review, 70(2), 725-772. | **D:** cohort<br>**S:** america<br>**V:**Natural disaster development<br>**I:** International development projects | Disaster development in the community |
| 9 | Nugrahandika W, Putri R. 2021 | **D:** Cross Secioal<br>**S:** Bantul residents<br>**V:**Emergency and Disaster Risk Management<br>**I:** Earth and Environment | Community-based disaster management |
| 10 | Pandey, CL (2019). Making communities disaster resilient: Challenges and prospects for community engagement in Nepal TT - Making communities disaster resilient. Disaster Prevention and Management, 28(1), 106–118. https://doi.org/http://dx.doi.org/10.1108/DPM-05-2018-0156 | **D:** literature review<br>**S:** 2 communities<br>**V:**Making communities resilient to disasters<br>**I:** CBDRM approach | The CBDRM approach motivates people to work together because they feel a sense of belonging to their community and recognize the benefits of their involvement in disaster mitigation and preparedness. |
| 11 | Pambudi, DI, & Ashari, A. (2019). Enhancing role of elementary school in developing sustainable disaster preparedness: a review with some examples from disaster-prone areas of Merapi. IOP Conference Series. Earth and Environmental Science, 271(1).<br>doi:http://dx.doi.org/10.1088/1755-1315/271/1/012016 |  |  |
| 12 | Putri, RP, Safitri, FN, Munir, S., & Endiyono, AH (2019). Basic Life Support Training Using Phantom Cardiopulmonary Resuscitation Media (Prejaru) Improves Basic Life Support Knowledge and Skills in Lay People. Emergency Journal. | **D: Cross Secioal**<br>**S:**<br>**V:**Emergency and Disaster Risk Management<br>**I:** training | Development of basic life support |
| 13 | Report, C. (2021). Disaster Preparedness in Neonatal Intensive Care Units. 139(5).<br>doi:10.1542/peds.2017-0507 | **D: Cohort**<br>**S:**<br>**V:**Emergency and Disaster Risk Management<br>**I:** training | Intensive care |
| 14 | Risk, D., & Framework, M. Emergency and Disaster Risk Management Framework. | **D: -**<br>**S: whole country**<br>**V:**Disaster Risk Management Framework.<br>**I:** module | disaster risk reduction to achieve inclusive sustainable development and the goals of the Sendai Framework. |
| 15 | Rahmayanti, KP, Azzahra, S., & Arnanda, NA (2021). | **D:** purposive sampling<br>**S:** Jakarta<br>**V:** Jakarta flood disaster<br>**I:** module | The people of Jakarta who are victims of floods |
| 16 | Rosya, A., & Dini, DP (2021). Regional early action in dealing with the threat of hydrometeorological disasters. (September). | **D:** purposive sampling<br>**S:**<br>**V:** threat of hydrometeorological disaster<br>**I:** module | Flood threat |
| 17 | Riama, NF, Sari, RF, Rahmayanti, H., Sulistya, W., & Nurrahmat, MH (2021). The Level of Public Acceptance to the Development of a Coastal Flooding Early Warning System in Jakarta. | **D:** purposive sampling<br>**S:jakarta people**<br>**V:**Development of Coastal Flood Early | In this study, we conclude that the level of public acceptance of the development of an early warning |
|  | Sustainability, 13(2), 566.<br>https://doi.org/http://dx.doi.org/10.3390/su13020566 | Warning System in Jakarta<br>**I**:questionnaire | system is very good as evidenced by the high mean scores of respondents' attitudes, |
| 18 | Saad, MFM, Latif, AA, & Othman, M. (2021). Agile information-based framework for flood management. IOP Conference Series. Materials Science and Engineering, 1088(1).<br>[doi:http://dx.doi.org/10.1088/1757-899X/1088/1/012048](http://dx.doi.org/10.1088/1757-899X/1088/1/012048) | **D**: purposive sampling<br>**S:jakarta people**<br>**V**:Development of Coastal Flood Early Warning System in Jakarta<br>**I**:questionnaire | In this study, we conclude that the level of public acceptance of the development of an early warning system is very good as evidenced by the high mean scores of respondents' attitudes, |
| 19 | Saddam, S., Lestanata, Y., Isnaini, I., Ihsan, I., Saoki, M., & Jafar, MUA (2019). TRAINING OF THE NATIONAL WATER SEARCH AND RESCUE (PP-SAR) FIRST AID TRAINING OF THE INDONESIAN RED CROSS VOLUNTEER CORPORATE AND HIGHER CORPORATE VOLUNTEER AROUND INDONESIA. JMM (Journal of Independent Society).<br>[doi:10.31764/jmm.v2i1.1339](https://doi.org/10.31764/jmm.v2i1.1339) |  |  |
| 20 | Schatten, M., & Tomi, I. (2017). A Review on Application Domains of Large-Scale Multiagents. (8537), 201-206. |  |  |
| 21 | Seda, G., & Parrish, JS (2019). Augmenting Critical Capacity in a Disaster. 35, 563-573.<br>[doi:10.1016/j.ccc.2019.06.007](https://doi.org/10.1016/j.ccc.2019.06.007) |  |  |
| 22 | Septiana, ME, Wardoyo, MAI, Praptiwi, NY, Ashari, ANS, Ashari, A., Susanti, NI, Jainudin, | **D**: Correlation<br>**S**:<br>**V**:Disaster education<br>**I**: local culture | efforts to support disaster management that have been carried |
|  | Latifah, F., & Nugrahagung, PP (2019). |  | out by the government. |
| 23 | SFDRR. (2021). Sendai Framework for Reduction. | **D:** -<br>**S:** whole country<br>**V:** sendai framework<br>**I:** module | Our mission is to provide leadership and support to accelerate global efforts in disaster risk reduction to achieve inclusive sustainable development and the goals of the Sendai Framework. |
| 24 | Sinta, D. (2021). Disaster literacy among young peatland farmers in Central Kalimantan. In (Vol. 249). Les Ulis: EDP Sciences. | **D:** purposive sampling<br>**S:** Kalimantan people<br>**V:** peatland farmers in Central Kalimantan<br>**I:** module | Disaster study in Kalimantan |
| 25 | Sorani, M., Tourani, S., Khankeh, HR, & Panahi, S. (2018). Prehospital Emergency Medical Services Challenges in. 6(6), 1-6. | **D:** cross sectional<br>**S:** -<br>**V:** Emergency Medical Services Challenges in<br>**I:** module | Treatment in the prehospital area |
| 26 | UNDRR. (2022). Strategic_Framework. | **D:**<br>**S:** whole country<br>**V:** sendai framework<br>**I:** module | leadership and support to accelerate global efforts in disaster risk reduction |
| 27 | Wirawan, CA, & Arsa, SAW (2020). | **D:** R and D<br>**S:** 50 nurses<br>**V:** handling cardiac arrest outside the hospital<br>**I:** BLS module | The basic android basic life support (BLS) application can increase the accuracy and speed of nurses in conducting BLS |
| 28 | Wright, N., F, L., Lapitan, JM, Kayano, R., Abrahams, J., Huda, Q., & Murray, V. (2020). | **D:** cohort<br>**S:** all countries<br>**V:** Emergency and Disaster Risk Management<br>**I:** SFDRR module | Overcoming these hurdles is important, and doing so will be a key marker of the success of the partnership over the next 10 years under the Sendai Framework. |
| 29 | Tam, G., Ying, E., & Chan, Y. (2019). Planning of a Health Emergency Disaster Risk Management Program for a Chinese Ethnic Minority Community. 1-12. doi:10.3390/ijerph16061046 | **D: cohort**<br>**S: Chinese people**<br>**V: Health Emergency Disaster Risk Management**<br>**I: module** | Early disaster management |
| 30 | Wanner, MST (2020). Drivers of Change in National Disaster Governance under the Hyogo Framework for Action. 8(4), 256-269. doi:10.17645/pag.v8i4.3062 | **D: cohort**<br>**S:-**<br>**V: Disaster Governance**<br>**I: module** | Application of the hyogo framework |
| 31 | Wisner, B. (2020). Five Years Beyond Sendai — Can We Get Beyond Frameworks ? International Journal of Disaster Risk Science, 11(2), 239-249. doi:10.1007/s13753-020-00263-0 | **D: cohort**<br>**S:-**<br>**V: International Journal of Disaster Risk Science**<br>**I: module** | Application of the sendai framework |
| 32 | Woman, JM (2019). Living Compound Marginality : Experiences of a. 1-26. | **D: cohort**<br>**S:-**<br>**V: Living Compound Marginality**<br>**I: module** |  |
| 33 | Ying, E., Chan, Y., Huang, Z., Siu, E., Lo, K., Kei, K., . . . Wong, S. Sociodemographic Predictors of Health Risk Perception, Attitude and Behavior Practices Associated with Health-Emergency Disaster Risk Management for Biological Hazards: The Case of COVID-19 Pandemic in Hong Kong, SAR China. (January 2020), 1-18. | **D: cohort**<br>**S: Chinese people**<br>**V: Attitude and Behavior Practices**<br>**I: module** | Community behavior in dealing with disasters |
| 34 | Zhang, L. (2021). Pre-Positioning Facility Location and Resource Allocation in Humanitarian Relief Operations Considering Deprivation Costs. Sustainability, | **D: cohort**<br>**S: Chinese people**<br>**V: Humanitarian Relief Operations** | Better development |
|  | 13(8), 4141-4141.<br>doi:http://dx.doi.org/10.3390/su13084141 | Considering<br>Deprivation Costs<br>I: module |  |

## discussion

### Pre Hospital

An important component of the national emergency care system is the prehospital emergency medical service(Duong et al., 2018). Community health centers as first-level health facilities must be equipped with basic emergency service capabilities to support an optimal health care system. The ability of nurses to perform cardiac massage or BLS is still below 50%(Wirawan & Arsa, 2020). The 2020 guidelines offer suggestions for increasing lay rescuer CPR rates, noting that currently less than 40% of non-hospitalized adults who experience cardiac arrest receive lay rescuer-initiated CPR prior to the arrival of emergency medical services.(Cpr & Ecc, 2020). The 2020 American Heart Association (AHA) Guidelines for Cardiopulmonary Resuscitation (CPR) and Emergency Cardiovascular Care provide a comprehensive review of evidence-based recommendations for resuscitation and emergencies.(Merchant et al., 2020)

Taken together, these frame works aim for a more complete agenda for action that includes health, development, humanitarian action, disaster risk management, and climate change adaptation.(Wright et al., 2020)

### Disaster

Disaster is an event or series of events that threaten and disrupt people’s lives and livelihoods caused by both natural and/or non-natural factors as well as human factors, resulting in human casualties, environmental damage, property losses, and psychological impacts. (BNPB, 2021)

Strengthening across sectors is very important for reducing disaster risk, which is continuously carried out continuously for community resilience in the disaster sector to issue resilience in disaster response(SFDRR, 2021)

### Type of disaster

1. Events caused by natural disasters are a series of natural events such as tsunamis, volcanic eruptions, landslides, floods, droughts, earthquakes.
2. A series of events that are not caused by nature are events in the form of failures due to modernization, and technology, epidemiology and diseases caused by epidemics.
3. Events caused by humans are called man-made disasters which include between groups due to disputes.
4. Disasters that are documented based on events and locations, types of disasters and resulting damage can be called disaster events.
5. Earthquakes are a series of vibrations or shocks that occur on the earth’s surface caused by the meeting of the earth’s plates, active faults, and active volcanoes or rock collapse.
6. Volcanic eruptions are part of a volcanic event known as an “eruption”. The dangers of volcanic eruptions can be in the form of hot clouds, incandescent ejections, heavy ash rain, lava, poison gas, tsunamis and lahars.
7. The word tsunami comes from the Japanese language which means waves or ocean waves (“tsu” means ocean, “nami” means waves). This event is the result of an earthquake from the ocean.
8. Stable soil can cause soil and rock movements that descend from this slope, which is called a landslide
9. The increase in the volume of water that causes the land to be submerged is a flood.
10. The addition of direct water discharge in large quantities and there are obstacles in river flow resulting in flash floods.
11. The need for water is a lot and there is water scarcity, while the need for water can still have an impact on the agricultural sector which leads to the economic factor of selling plants or plants that are traded.
12. A house or settlement that is hit by a fire is a series of fire events that cause casualties or losses
13. Forests with land that are engulfed by fire, resulting in forest and land damage that cause economic losses and or environmental values. Forest and land fires often cause smoke disasters that can disrupt the activities and health of the surrounding community.
14. The emergence of very strong winds with circular movements resembling a spiral can reach speeds of up to 50 km / h, this event is called a hurricane which will disappear within 3-5 minutes (BNPB, 2020)

## Conclusion

1. The development of a disaster education model is very important in the initial handling of emergencies in the pre-hospital area in disaster risk management
2. Development of a disaster education model in the initial handling of emergencies in the pre-hospital area involving multi-agent bases or lay people in disaster risk reduction

## Data Availability

This article uses review literature so does not use clinical trials
all data from BNPB or National agencies and disaster management

https://bnpb.go.id/definisi-bencana

## Conflict of Interest Disclosure Form

All authors need to complete and submit this form when submitting a manuscript. Disclosures and signatures from all authors on one form is preferred.

*Please note that a conflict of interest statement is published with each paper and must be inserted in your text document right before the reference list*.

I/we certify that there is no actual or potential conflict of interest in relation to this article.

(Please print names)

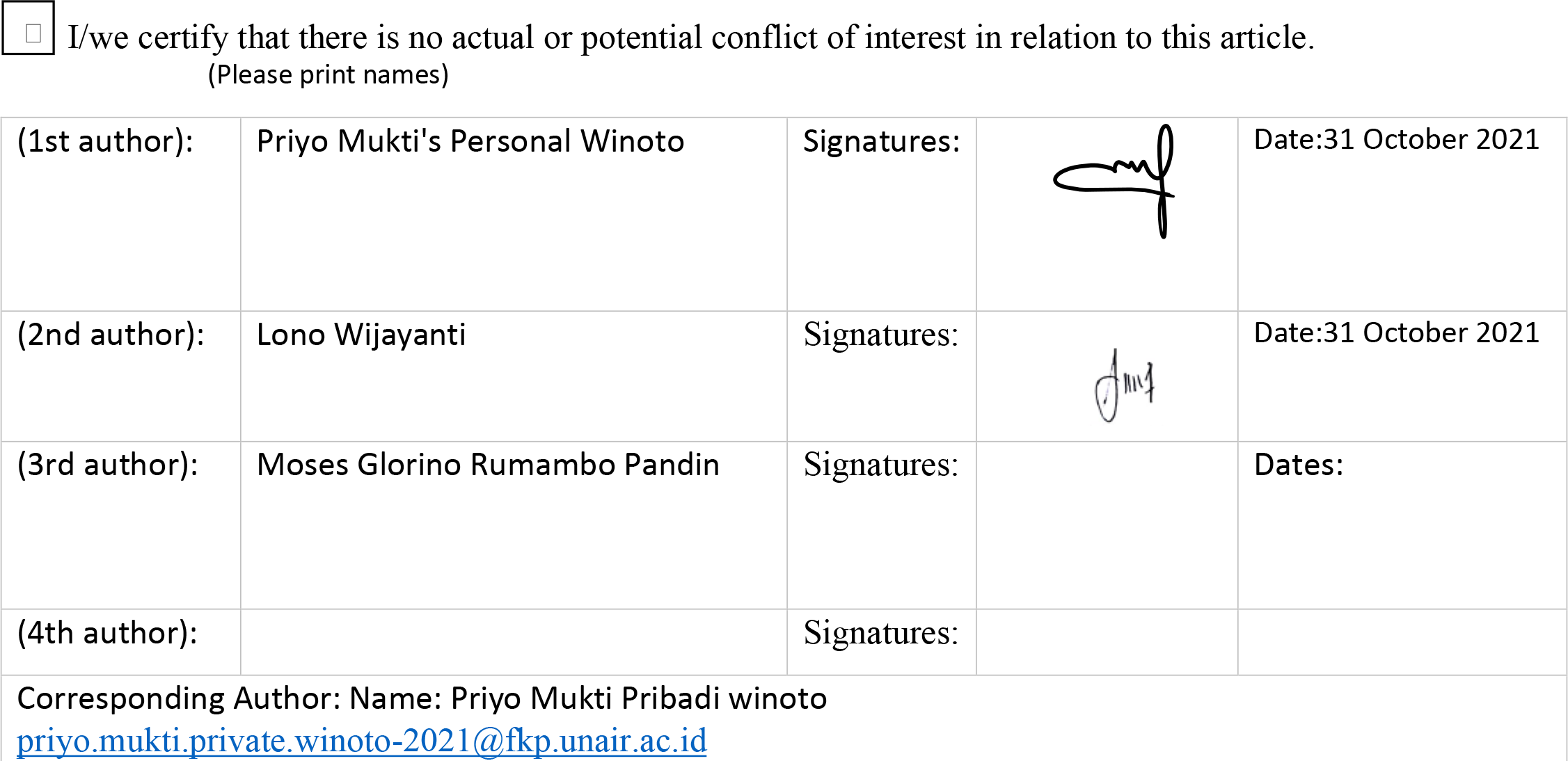

|  |  |  |  |  |
| --- | --- | --- | --- | --- |
| (1st author): | Priyo Mukti's Personal Winoto | Signatures: |  | Date:31 October 2021 |
| (2nd author): | Lono Wijayanti | Signatures: |  | Date:31 October 2021 |
| (3rd author): | Moses Glorino Rumambo Pandin | Signatures: |  | Dates: |
| (4th author): |  | Signatures: |  |  |

